# Estimating and forecasting COVID-19 attack rates and mortality

**DOI:** 10.1101/2020.05.11.20097972

**Authors:** David I. Ketcheson, Hernando C. Ombao, Paula Moraga, Tarig Ballal, Carlos M. Duarte

## Abstract

We describe a model for estimating past and current infections as well as future deaths due to the ongoing COVID-19 pandemic. The model does not use confirmed case numbers and is based instead on recorded numbers of deaths and on the age-specific population distribution. A regularized deconvolution technique is used to infer past infections from recorded deaths. Forecasting is based on a compartmental SIR-type model, combined with a probability distribution for the time from infection to death. The effect of non-pharmaceutical interventions (NPIs) is modelled empirically, based on recent trends in the death rate. The model can also be used to study counterfactual scenarios based on hypothetical NPI policies.

## 1 Introduction

The COVID-19 pandemic progressively propagated from Wuhan, China, where it initiated in December 2019 to achieve pandemic status affecting 173 nations of 195 nations worldwide by April 2020^1^. In many affected nations, COVID-19 has become a leading cause of mortality, with eight European nations (Belgium, UK, Spain, Italy, The Netherlands, San Marino, Andorra and France) topping the accumulated fatality rate, exceeding 300 individuals per million inhabitants ^2^.

While effective therapies and vaccines are being developed, confinement emerges, as in the Spanish flu pandemic a century ago [26], as the leading policy to mitigate the impacts, by reducing the peak number of infections, hospitalizations and fatalities [21, 20]. However, strict confinement measures to mitigate against COVID-19, as recommended by WHO, are being disputed for their impacts on the economy and jobs. The development of cost-benefit analysis of confinement measures requires reliable projections of both the costs to the economy and the health cost (Mubayi et al. 2010), ultimately dependent on the fatality rate.

Exit strategies that attempt to minimize both public health and economic impacts depend critically on reliable models for both. The fundamentals of analytical models to predict disease spread were laid a century ago (e.g. [14]), allowing models of the health risks associated with COVID-19 to emerge rapidly [11, 13, 17]). The performance of these models depends critically on the quality of the initial data referring to the population affected (total population size, people diagnosed and deaths attributed to COVID-19 over time), the parameters specific to the COVID-19 propagation and impact (e.g. R0, Fatality rate), and the impact of mitigation measures on those parameters [22, 24]). As COVID-19 spreads to many countries and other situations, such as vessels with large confined populations, multiple series of data are emerging that offer opportunities to improve the assessment of key parameters. Moreover, as the effort and resolution of testing varies over time, it has become evident that the number of diagnosed cases is unreliable as it depends on effort and particular kits used, which vary among and within nations. Although the number of deaths reported may also be underestimated, as COVID-19 deaths may be undetected or confounded with other causes, it provides a minimum estimate that is reported in a relatively consistent manner across many nations, following WHO guidelines, which require fatalities to be listed only where the disease caused, or is assumed to have caused, or contributed to death.^3^

### 1.1 Modeling approach

Most available models make us of confirmed cases as the primary input or calibration; for example [1, 3, 4, 5, 6, 7]. Confirmed case numbers drastically underestimate the real number of infections; furthermore, the ratio of real infections to confirmed cases varies greatly both in time and from region to region, due to differences in testing and in reporting policies (see for example Figure 3 herein). Some models also ignore the fundamentals of epidemiology and are based merely on curve-fitting; for example [2, 3].

Here we describe a model for inferring the past and present attack rate, and predicting the death toll over time due to the ongoing COVID-19 pandemic. The model is based on recorded numbers of deaths and COVID-19 parameters derived from estimates in the literature; it makes predictions of daily numbers of new infected persons and deaths. The effect of non-pharmaceutical interventions (NPIs) such as school closures, prohibition of public events, and restrictions on internal movement is modelled empirically, based on recent trends in the death rate reported for a number of nations and of states in the USA following different levels of normative intervention effected in these case studies. The model can also be used to study counterfactual scenarios based on hypothetical NPI scenarios, and, therefore, be used to explore different proposed policies.

Highly sophisticated epidemiological models involving, for instance, spatial resolution and networks of social connections, can be of great use in making detailed predictions, provided that the relevant parameters are adequately known. This is not yet the case for COVID-19, given that estimates of the basic reproduction number itself differ substantially [31]. As a result, current predictions of COVID-19 infections and deaths differ by orders of magnitude [10]. Most of those predictions are based on confirmed case numbers, which are known to be highly inaccurate. Given these large uncertainties, we propose that a straightforward model based on relatively well-established inputs may be more useful than a complex model requiring poorly-constrained or highly inaccurate data and parameters. The present work is based on one of the simplest epidemiological models available, but with a focus on using the most accurate available inputs. The technique used herein to infer infections from recorded deaths could also be used to provide more accurate input to more sophisticated models, in place of confirmed case numbers.

The model algorithm is described in Section 2. The model data sources and values of model parameters are given in Section 3. In Section 4 we study several model results, including predictions for a counterfactual scenario showing what might have happened if nothing was done to slow the spread of COVID-19. Finally, in Section 5 we discuss the main sources of error and uncertainty in the model.

The model code, along with scripts to generate the figures and tables in this paper, are available.^4^

## 2 Model algorithm

The central element of the forecasting algorithm is based on the classic SIR model [14]:

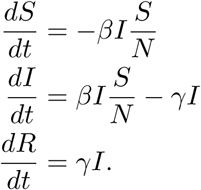

Here *S*(*t*)*,I*(*t*)*,R*(*t*) represent the susceptible, infected, and removed (recovered or deceased) fractions of the population, respectively. To account for NPIs, we add a time-dependent intervention parameter *q*(*t*), which models the change in the contact rate *β* due to social distancing, school and work closures, and other such measures. This model takes the following form [15]:

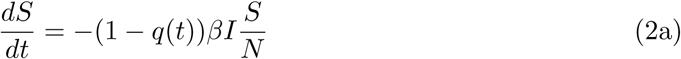

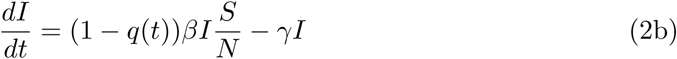

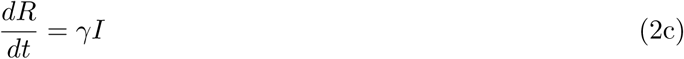

A value *q*(*t*) = 0 corresponds to no intervention, while *q*(*t*) = 1 corresponds to a complete elimination of contact with infected individuals. This parameter can model both population-wide measures and those specifically targeting infected or suspected individuals. The time-dependent reproduction number is a measure of transmissibility that denotes the average number of secondary cases caused by an infected individual in a fully-susceptible population and can be expressed as

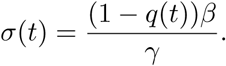

In the forecast model, *β* and *γ* (which are primarily properties of the disease) are fixed for all countries and all time, but *q*(*t*) (which is primarily an effect of human actions) is time-dependent and different for each country.^5^

Many further extensions of the SIR model exist, in which other groups, such as those under quarantine are directly modeled. Our rationale for using the model (2) is twofold. First, due to the prevalence of asymptomatic cases and broad social-distancing measures, the dominant effect of intervention is through general reduction in the contact rate rather than quarantining of a small group. Second, the existing uncertainties in key parameters for this still-developing pandemic are likely to be much greater than the differences between various refinements of the model.

### 2.1 Model structure

Let *j* indicate the date, where for convenience *j* = 0 denotes the present day.

- *S_j_*: Susceptible individuals on day *j*
- *i_j_*: Newly infected individuals on day *j*; *i_j_* = *S_j_*_−1_ − *S_j_*
- *I_j_*: Currently infectious individuals on day *j*
- *D_j_*: Cumulative deaths up to day *j*
- *d_j_*: New deaths on day *j*; *d_j_* = *D_j_ − D_j_*_−1_
- *R_j_*: Cumulative removed (including both recovered and deceased) individuals on day *j*.

As depicted in Figure 1, the forecast model involves three main parts:

1. Inference of real past infection rates based on recorded numbers of deaths up to a few weeks in the past (to avoid bias due to cases that have not yet died but will do so in the future).
2. Simultaneous modeling and fitting of data over a recent time period approximately equal to the mean time from infection to death.
3. Forecasting future values based on a hypothetical level of intervention.

**Figure 1:**
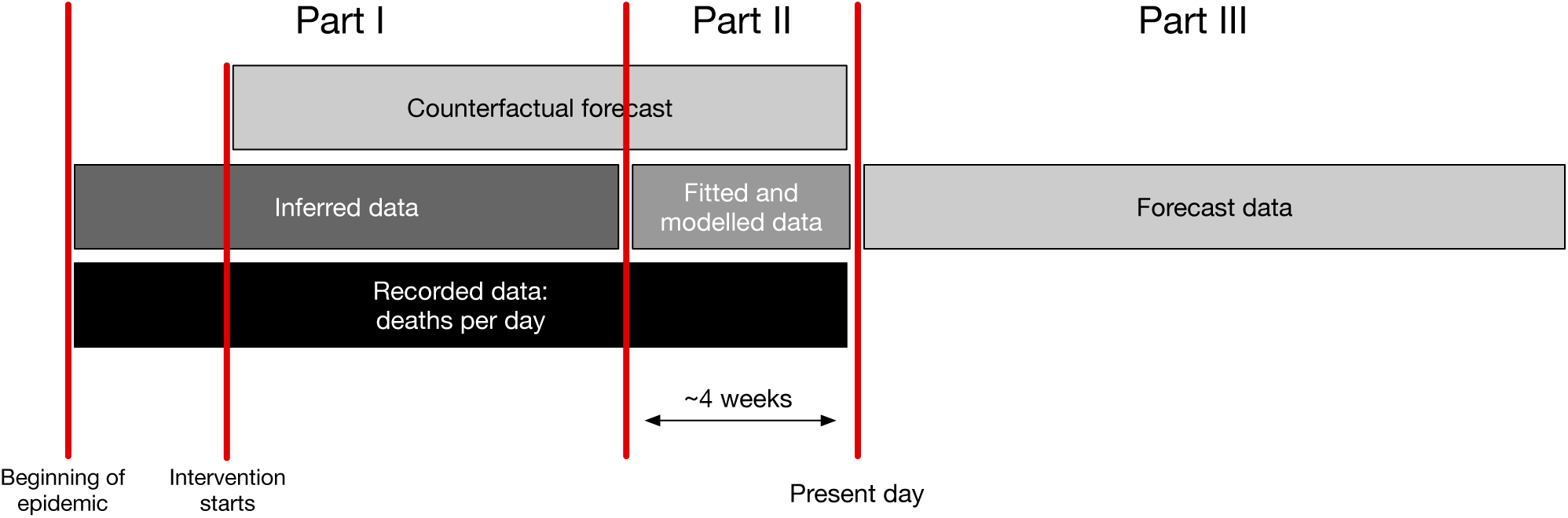
Organization of the elements of the model.

Each of these parts is described in the following sections.

### 2.2 Part I: Inference of past attack rate

In this part we infer the number of newly infected each day, *i_j_*, from the number of deaths, *d_j_*. Let *F* denote the mean infection fatality ratio *F* for a population (see Section 3). Based on published estimates we approximate the distribution (among patients who die) of time from infection to death by a discretized gamma distribution with mean 14 days and standard deviation 6.01 days, plotted in Figure 2. Let *P_k_* denote the probability that a newly infected person dies *k* days after infection. This distribution is normalized based on *F* so that

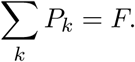

**Figure 2:**
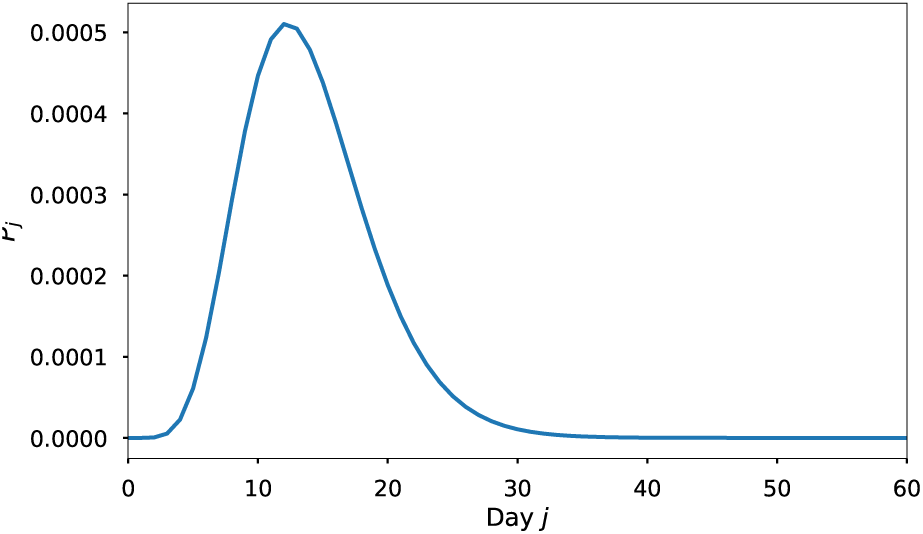
Probability distribution for time from infection to death.

The daily deaths *d_j_* are given by the discrete convolution of the new infections *i_j_* with the distribution *P*:

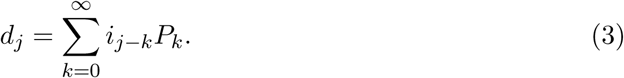

The new infections can thus be found by a discrete deconvolution, which is ill-conditioned (i.e., the result is highly sensitive to small changes in the data). Due to non-smoothness of real-world recorded death rates, the deconvolved infection rate is typically highly oscillatory and includes negative values.

#### 2.2.1 Deconvolution algorithm for inferring infections from deaths

In order to obtain realistic values, we first smooth the daily death values using a 3-day moving average and then use a non-negative least squares approach with regularization [19]. To apply this approach to data recorded over *J* days, we write (3) in the matrix-vector form

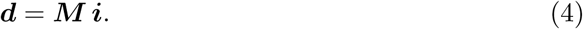

Here d = [*d_j_*], *j* = 1, …, *J*; *M* is a *J* × *J* Toeplitz matrix with a first column given by *P* = [*P_k_*], *k* = 1, …, *J*; and *i* = [*i_j_*], *j* = 1, …, *J*. An estimate 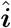 of the daily infections can be obtained by pursuing the following optimization [9, 27]:

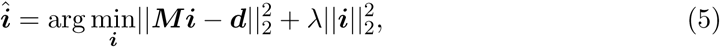

where ||·||_2_ is the Euclidean norm and λ is a positive real-valued regularization parameter.

A standard non-negative least-squares solver can be used to perform the minimization in (5). We have found that this approach gives satisfactory results except during the very earliest stage of the epidemic, when it tends to significantly underestimate the number of infections. This effect can be attributed to the fact that the probability vector ***P*** represents the average case, which introduces bias by underestimating the contributions from smaller daily infection occurrences. To remedy this, we apply the following perturbation based technique. We perform many random trials of generating the model (4) based on a perturbed version of the probability vector ***P***. Namely, we generate

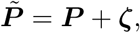

where ζ is a random noise vector. Then we use 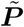 to generate a convolution matrix 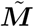 which is used to replace the matrix ***M*** and obtain an estimate of *i* using (5). The process is repeated for a sufficient number of trials. The final estimate of *i* is formed by taking the average of the estimates obtained from all trials.

This deconvolution gives values of *i_j_* up to the present day, but the most recent values are not accurate since they would be influenced significantly by unknown future values of deaths. We therefore use these values only up to *m* = 21 days in the past; this ensures that for all dates of inference, at least 90% of the resulting deaths would have occurred by the present date.

Removal is modeled by a Poisson process in which the probability of recovery in a given one-day period is *γ*. The cumulative number of recovered individuals at day *j* is then computed as

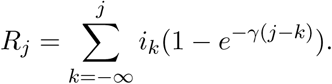

The number of actively infected persons on day *j* is then

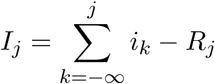

The number of susceptibles on day *j* is

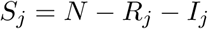

### 2.3 Part II: fitting recent past data

In order to estimate the infection rate up to the present date, we model the most recent *m* days using the SIR model (2). We fit the recent data by minimizing a weighted sum of squares of the difference between the logarithm of the predicted and observed deaths, where the weighting emphasizes the most recent values:

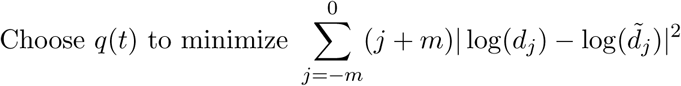

where the modeled number of deaths is

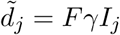

with *I_j_* given by the solution of (2) subject to the initial susceptible and infected fractions (*S***_−_***_m_, I***_−_***_m_*), obtained from the inference model in part I. Two fits are performed; in the first, intervention is assumed to be constant:

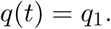

In the second the intervention is assumed to be a linear function:

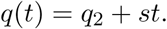

The fit parameters are constrained so that *q*(*t*) remains in the interval [0,1]. To reduce the sensitivity to noise in the data, this fit includes a penalty proportional to the magnitude of the slope. In both fits, the fit is weighted to be more sensitive to the most recent data. We have found that the constant fit *q*_1_ is more robust but the linear fit is better at adapting to recent changes in intervention.

### 2.4 Part 3: forecasting

From Part 2 we obtain estimate values (*S*_0_, *I*_0_*, R*_0_) at the present day. These are combined with a hypothesized future intervention effectiveness *q*(*t*) and fed into the augmented SIR model (2) to predict future dynamics of the pandemic. The model is not intended to predict the long-term intervention effectiveness, which is largely a matter of government policy. In the short term, as a compromise between robustness and flexibility, we estimate the intervention effectiveness by the average of the two fits from part 2, evaluated at the present day:

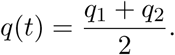

Forecasts can also be generated using some assumed future intervention policy that changes in time.

## 3 Parameter values and data sources

Here we describe the main inputs and parameters of the model, along with the values used. For both inputs and parameters, the model can easily be used with other values than the specific sets chosen here. For instance, it might be useful to replace the officially reported COVID-19 deaths with excess mortality data.

### 3.1 Data

The primary model input is the number of confirmed COVID-19 deaths per day in a given region (a country or US state) since the start of the pandemic. The recorded deaths are obtained from:

- For countries: https://github.com/CSSEGISandData
- For states: https://github.com/nytimes

Because death counts are noisy in practice, we smooth these time series by using a 3-day moving average. In order to compute the mean IFR, we use population data from the UN [8].

### 3.2 Parameters

Next we discuss the key parameters and give the values used in the model.

- Mean infectious period: 14 days [18, 28]. This enters the model through *γ* = 1/14.
- Basic reproduction number *σ*_0_: 3.0 (2.0-4.0). This enters the model through *β* = *σ*_0_*γ*. Early estimates of the basic reproduction number were low, with some even less than two [31], but some more recent estimates are closer to four [25].
- The infection fatality ratio *F*. This is the fraction of infected persons that die from COVID-19. Since the IFR for COVID-19 is known to vary strongly with age, we use age-specific IFR estimates from [29]. These are combined with UN demographic data for each country [8] to determine an effective IFR, where we assume a uniform attack rate across age groups:

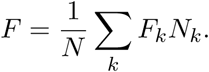 Here *F_k_* is the IFR for age group *k* and *N_k_* is the population in age group *k*. For regions in which we lack detailed demographic data, we use a value of 0.6% based on estimates from [23, 31, 29].

## 4 Exploration of model outputs

### 4.1 Real vs. confirmed infections

We can use the inferred infection rates from Part I above to estimate the degree to which cases in a given region are under-reported. Table 1 shows this comparison for the date of March 9th. We compare the values from our model with those of [16], which used a very different approach.

**Table 1:** Comparison of reported and estimated cumulative infections for several countries, as of March 9th. The last column shows, for comparison, estimated under-reporting from a different model in [16].

| Region | Reported | Estimated | Difference | Ref. [16] |
| --- | --- | --- | --- | --- |
| China | 80,763 | 409,353 | 328,590 | 12-89 million |
| Italy | 8,889 | 381,011 | 372,122 | 30,223 |
| Iran | 7,248 | 398,766 | 391,518 | 266,213 |
| South Korea | 7,416 | 30,111 | 22,695 | 18,809 |
| France | 1,369 | 46,214 | 44,845 | 7,931 |
| Spain | 1,140 | 121,806 | 120,666 | 87,405 |
| Germany | 1,215 | 5,180 | 3,965 | 2,277 |
| US | 675 | 40,341 | 39,666 | 1.21 million |

In Figure 3 we compare confirmed cases with inferred numbers of infections. The values plotted are the daily increase in each number. For Spain, Italy, and the USA we observe a similar trend, with a huge gap (on the order of a 100x ratio) between the real and confirmed values, which later narrows to a factor of four or five. In contrast, South Korea (where the pandemic was rapidly controlled) and Saudi Arabia (where the pandemic has grown at a relatively slower rate) show a much higher detection rate early on, but similar rates later.

**Figure 3:**
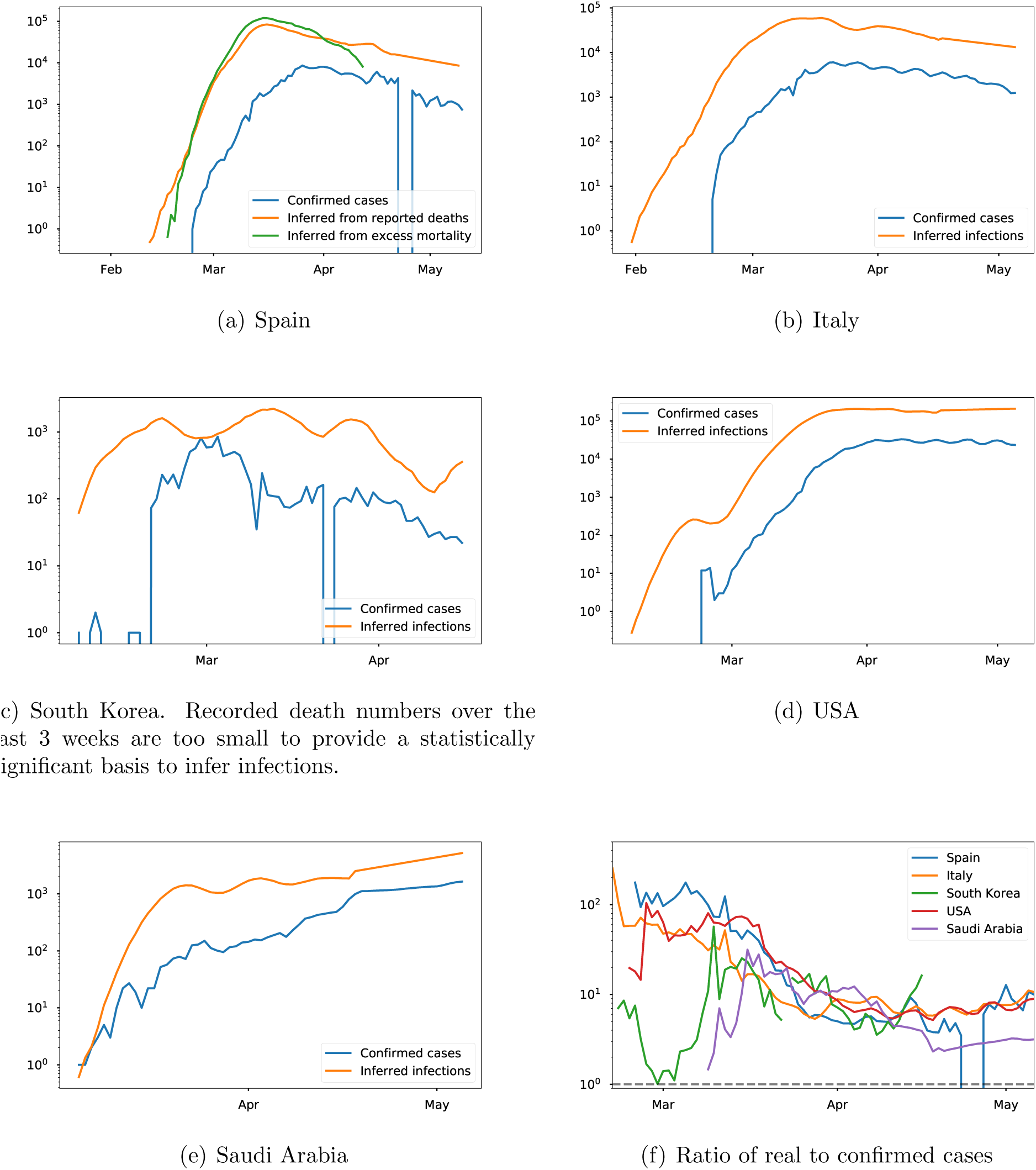
Inferred infections vs. confirmed infections.

Note that the confirmed case counts include some negative values, due to after-the-fact revisions made by some reporting organizations. The avoidance of this kind of artifact is another benefit of using recorded deaths as the basis (which so far have been much less affected by such artifacts) for modeling. Still, confirmed deaths are not completely accurate and may be supplemented by statistical estimations of excess mortality. In Figure 3 we show for Spain only the rate of infection inferred based on excess mortality^6^. As one might expect, it is more difficult to capture the early rise in infection as the deviation from expected mortality is at first not statistically significant. On the other hand, when the infection rate is high the excess mortality gives a more accurate accounting and implies higher rates of infection.

### 4.2 Estimating current immunity

Next we investigate the fraction of the population estimated to have antibodies (i.e., all of those who have been infected). Based on the behavior of other viruses, it is reasonable to expect that this portion of the population has some degree of immunity at present. Results for the 10 most-affected countries and the 10 most-affected US states are shown in Figure 4. Currently-infected individuals are included in this count.

**Figure 4:**
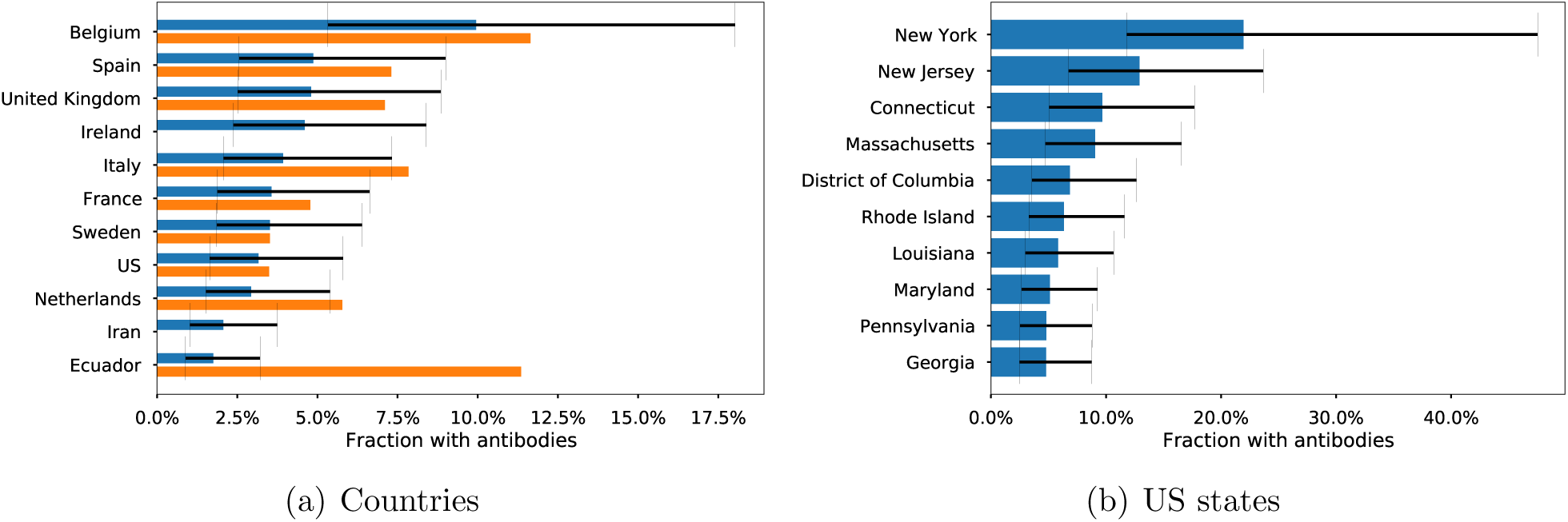
Estimated fraction of population with antibodies, showing the 10 countries and US states with the highest proportion of antibodies as of May 7th. Expected values and upper/lower bounds are based on infection fatality rates from [29]. Note the different horizontal scales. For countries, the orange bars show expected value corrected based on the ratio of excess mortality to recorded COVID-19 deaths. These numbers are not complete and are not available from all sources, but give some idea of how underreporting affects the estimates.

We remark that the values given are no more accurate than the total confirmed deaths for each region. Since excess mortality records suggest that deaths are substantially under-reported, we expect that actual antibody levels are significantly higher than what is computed here. This also means that the most hard-hit regions might not even appear in these figures if they have greatly under-reported fatalities.

Nevertheless, the values obtained here are broadly in agreement with rough serological surveys that have been conducted recently, indicating for instance that around 3% of the Netherlands and 15% of New York residents have antibodies. While these surveys did not necessarily adhere to scientific standards of sampling, the general agreement gives some confidence in these estimates.

### 4.3 Estimating intervention effectiveness

Here we try to infer values of the intervention effectiveness parameter *q*(*t*) based on the recorded data. The inference model above gives an estimate *i* of the number of new infections on day *j*, as well as estimates of *S_j_, I_j_, R_j_*. According to the SIR model with intervention (2), the number of new infections each day should be approximately

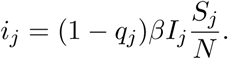

Thus

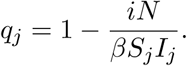

Some estimates of *q*(*t*) are plotted in Figure 5.

**Figure 5:**
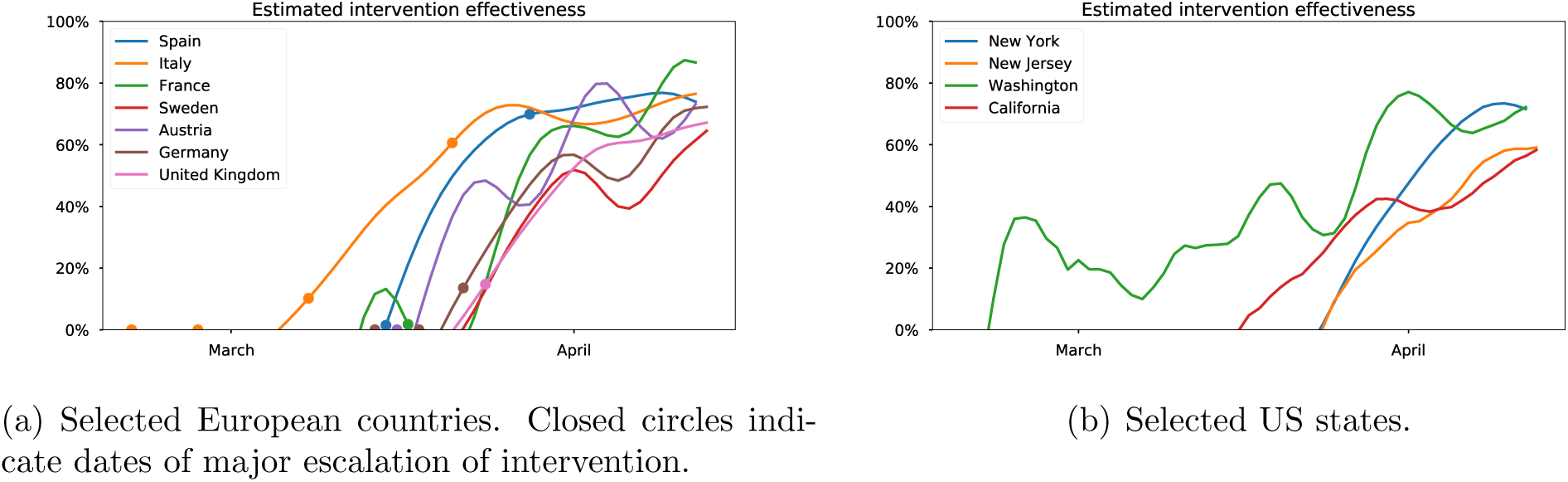
Estimated intervention effectiveness over time.

### 4.4 Comparison of forecasts with data

The model should not be viewed as a tool for giving absolute predictions of future infections and deaths. Rather, it provides predictions *conditioned* on an assumed future intervention effectiveness. We have equipped the model with a way to empirically estimate the intervention effectiveness q over the last 3 weeks, and a natural approach to prediction is to assume that the same value of q will apply in the future while interventions are maintained. This can also be used as a way to validate the model, by applying the model to a restricted data set in which the most recent values are omitted, and then *predicting* values for that recent time period. We expect this to give accurate results if there has been no policy change in the last few weeks. If intervention has changed significantly in the last few weeks, the results will be less accurate.

In Figure 6 we show results of this approach. We have omitted the last 14 days of data. We have intentionally chosen a difficult period for prediction, just after the death rate peaked in Spain and Italy.

**Figure 6:**
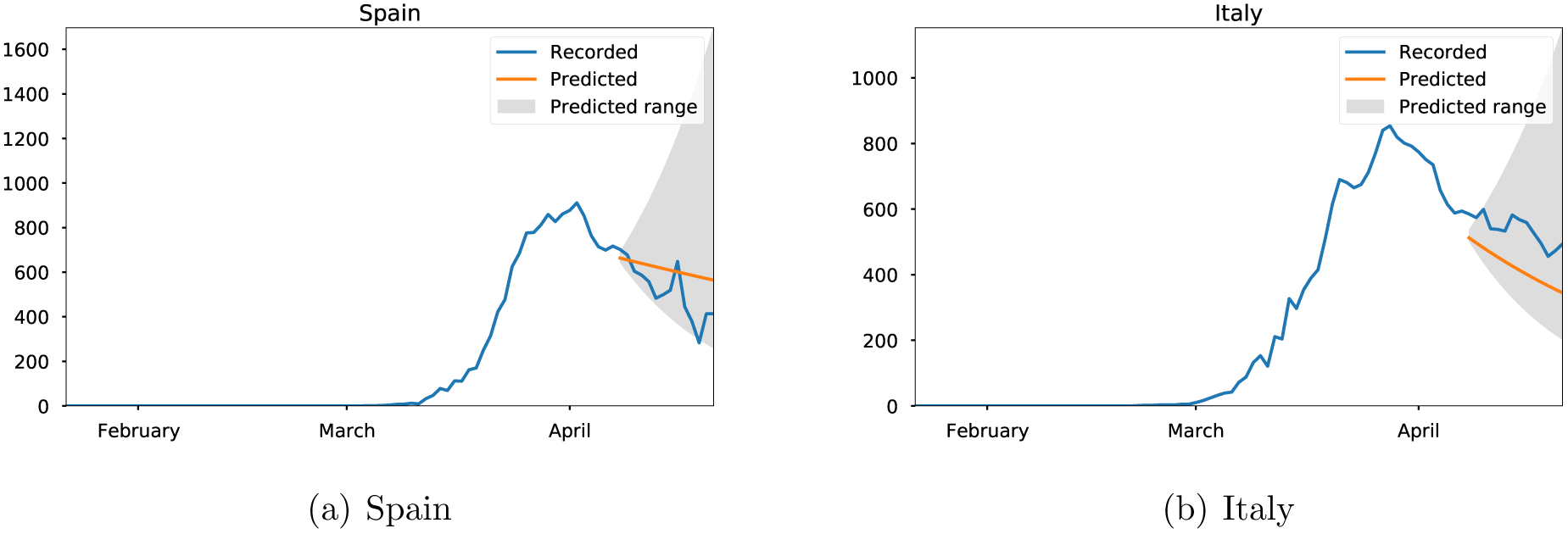
Predicted values obtained by omitting the most recent 14 days of data, compared with recorded values for daily deaths.

### 4.5 Modeling a no-intervention scenario

The model can also be used to generate a hypothetical scenario starting at some time in the past. The main purpose of this feature is to examine how the epidemic might have progressed in a given region in the absence of intervention. This involves the following steps:

1. Choose a starting date for the model. This should be a date before when intervention started, but after a statistically significant number of deaths had occurred. For some regions (those that are very small or where the pandemic started very late) these two requirements may be contradictory.
2. Determine initial numbers of susceptible, infected, and recovered for the starting date. These are determined using the inference model described in Section 2.2.
3. Solve the initial value problem consisting of the SIR model (1) (without intervention), from the chosen starting date.

Estimated deaths in the no-intervention scenario are compared with actual deaths in Figure 7 and Table 2.

**Figure 7:**
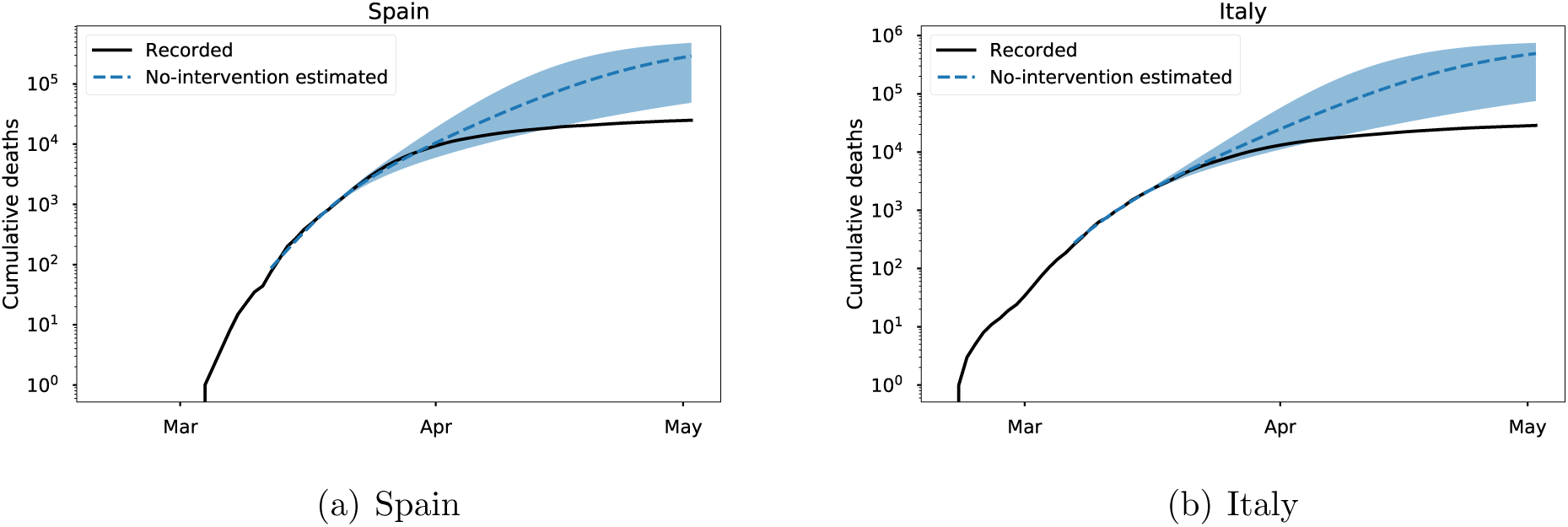
Estimated deaths in the absence of intervention, compared with recorded deaths.

**Table 2:** Estimation of deaths averted as of May 4th, 2020, due to intervention. The start date for each simulation is based on when the epidemic began to grow and when nationwide intervention was put in place. The values in parentheses represent the Bayesian credible interval assuming *σ_0_* ∈ [2.0, 4.0].

| Region | Starting date | Difference in estimated deaths |
| --- | --- | --- |
| Austria | 03-16-2020 | 10956 (796-48759) |
| Belgium | 03-20-2020 | 34889 (554-82118) |
| Denmark | 03-18-2020 | 7551 (651-30487) |
| France | 03-17-2020 | 191034 (9088-502624) |
| Germany | 03-14-2020 | 47879 (0-361409) |
| Italy | 03-07-2020 | 463653 (46781-718723) |
| Norway | 03-24-2020 | 3106 (441-13621) |
| Spain | 03-15-2020 | 282825 (36757-453079) |
| Switzerland | 03-20-2020 | 25562 (3361-61360) |
| United Kingdom | 03-24-2020 | 164636 (15864-400730) |

## 5 Sources of error and uncertainty

The quote from statistician G.E.P. Box that “all models are wrong, but some models are useful” is certainly applicable here. The difficulty of modeling a pandemic of a new disease while it is ongoing is high, and, given the uncertainties involved, all results should be taken as rough estimates. The primary sources of error and uncertainty include:

- Uncertainty in the key parameters *β, γ*, *F*. Published scientific estimates of each of these values vary by at least a factor of 2, and two of these parameters enter into the exponent of the exponential growth of the pandemic. Therefore, even small uncertainties in these parameters can lead to very large uncertainties in the model outputs.
- Inaccuracies in the input data. Our model intentionally disregards confirmed case numbers because they are a very poor indicator of real infections due inadequate testing and prevalence of mild or asymptomatic cases, as well as non-canonical testing standards [30] and false negatives depending on the diagnostic method used [16]. Numbers of deaths are typically much more accurate, but still contain substantial errors.
- Inaccurate model assumptions. The SIR model is one of the simplest epidemiological models. One of its key assumptions is that of homogeneous mixing among the population – i.e., that any two individuals are equally likely to have contact. This is of course far from true in the real world. More detailed models involving spatial structure and human mobility networks can partially remedy this deficiency. We have stuck to the SIR model so far because it seems that the parameter uncertainties already listed above – which will impact any epidemiological model – may be more significant than this modeling error.
- Modeling the impact of intervention. Other models, such as that of [12], have attempted to explicitly model the effect of officially-announced intervention policies, lowering the contact rate by some amount starting when the policy is put in place. This requires an inherently uncertain estimation of the quantitative impact of a given policy, which will certainly vary in time and between different societies. To avoid this, we use an empirical assessment of intervention, choosing a contact rate that reproduces the data. This contact rate is somewhat sensitive to the noisy daily death rate data. Additionally, the model is slow to adapt to changes in intervention policy, since their effects do not show up in the death rate until at least 2-3 weeks after they are implemented.
- Assumptions about future policy. Our primary forecast assumes that the current level of intervention will be maintained over the entire forecast period. For long-range forecasts, this is very unlikely. The model is not intended to show what will likely happen in reality, since future intervention policy is a matter of political choices and not susceptible to detailed mathematical modeling. Rather, the forecast values should be interpreted as showing what will likely happen under a specified intervention policy.

## Data Availability

Code and data are available from https://github.com/ketch/covid_forecasting.

## Footnotes

1 WHO, https://www.who.int

2 WHO, https://www.who.int, accessed 6 May, 2020

3 https://www.who.int/classifications/icd/Guidelines_Cause_of_Death_COVID-19.pdf

4 https://github.com/ketch/covid_forecasting

5 Can we also connect the (1 − *q*(*t*))*β*/*γ* as the effective reproduction rate at time *t* which takes into account the impact of an intervention at time *t*? This is now done.

6 Obtained from https://momo.isciii.es/public/momo/data.AccessedMay11th.

